# Early Prediction of Alzheimer’s Disease and Related Dementias Using Electronic Health Records

**DOI:** 10.1101/2020.06.13.20130401

**Authors:** Xi Yang, Qian Li, Yonghui Wu, Jiang Bian, Tianchen Lyu, Yi Guo, David Marra, Amber Miller, Elizabeth Shenkman, Demetrius Maraganore

## Abstract

Alzheimer’s disease (AD) and AD-related dementias (ADRD) are a class of neurodegenerative diseases affecting about 5.7 million Americans. There is no cure for AD/ADRD. Current interventions have modest effects and focus on attenuating cognitive impairment. Detection of patients at high risk of AD/ADRD is crucial for timely interventions to modify risk factors and primarily prevent cognitive decline and dementia, and thus to enhance the quality of life and reduce health care costs. This study seeks to investigate both knowledge-driven (where domain experts identify useful features) and data-driven (where machine learning models select useful features among all available data elements) approaches for AD/ADRD early prediction using real-world electronic health records (EHR) data from the University of Florida (UF) Health system. We identified a cohort of 59,799 patients and examined four widely used machine learning algorithms following a standard case-control study. We also examined the early prediction of AD/ADRD using patient information 0-years, 1-year, 3-years, and 5-years before the disease onset date. The experimental results showed that models based on the Gradient Boosting Trees (GBT) achieved the best performance for the data-driven approach and the Random Forests (RF) achieved the best performance for the knowledge-driven approach. Among all models, GBT using a data-driven approach achieved the best area under the curve (AUC) score of 0.7976, 0.7192, 0.6985, and 0.6798 for 0, 1, 3, 5-years prediction, respectively. We also examined the top features identified by the machine learning models and compared them with the knowledge-driven features identified by domain experts. Our study demonstrated the feasibility of using electronic health records for the early prediction of AD/ADRD and discovered potential challenges for future investigations.

## Introduction

Alzheimer’s disease (AD) and AD-related dementias (ADRD), a class of complicated neurodegenerative disorders that result in short-term memory lapses and lead to loss of bodily function and death^1^, pose a significant healthcare burden in the United States (US). In 2019, it was estimated that approximately 5.8 million Americans lived with AD/ADRD, where 97% of them are 65 years or older^2,3^. By 2050, the number of people living with AD/ADRD is expected to grow to 13.8 million, fueled by the aging baby boomers^4^. A total number of 121,404 AD/ADRD-related deaths were recorded in 2017, making AD the 6th leading cause of death for all ages and the 5th leading cause of death among people 65 years or older in the US^2^. Nevertheless, there is still no cure and no effective treatment for AD despite decades of investment^5–7^. The quality of life (QoL) of AD/ADRD patients is gradually diminished, and caring for AD/ADRD imposes a tremendous emotional and financial burden on patients, family caregivers, communities, and healthcare systems^8^. In 2019, the total cost of care for AD/ADRD patients was estimated at $ 290 billion to the healthcare system and 18.5 billion hours of informal (unpaid) assistance from the caregivers^2,9^. Drug development for AD/ADRD has faced enormous difficulty. From 2002 to 2012, over 99% of the AD/ADRD-related clinical trials failed; there were only 4 drugs approved for alleviating the symptoms of AD/ADRD since 1998 (and none for neuroprotection or neurorestoration)^10^. One reason for the high failure rate is that interventions given to the patients are too late (e.g., the trail of Bapineuzumab)^5^ as the neurodegenerative process of AD/ADRD starts years before the onset of clinical symptoms and the diagnosis. However, many of the risk factors are modifiable. Therefore, methods of identifying patients at high-risk of AD/ADRD before symptom onset are needed^11^.

Methods of predicting diseases have included the use of patients’ historical information for different time windows before the disease onset. Scientists have explored, for AD/ADRD prediction, various sources of patients’ data including medical images, patient claims, genomics, and electronic health records (EHRs) ^12,13^. Both statistical models and machine learning models have been explored extensively^14^. Many previous studies have investigated neuroimaging resources such as the comprehensive imaging-based data resource provided by the Alzheimer’s Disease Neuroimaging Initiative (ADNI)^15,16^. Previous works using neuroimaging data demonstrated good performance^17^, however, most of them focused on prediction of the conversion from mild cognitive impairment (MCI) to AD/ADRD^18–20^. In clinical practice, doctors do not employ neuroimaging procedures to predict AD/ADRD as the indication is not established and as the costs are prohibitive^21,22^. With the rapid adoption of EHR systems in hospitals, longitudinal, routinely collected patient care data has become available for clinical research^23,24^. Risk factors for cognitive decline and dementia (including ADRD), that are well supported in the literature (e.g., traumatic brain injury, obesity, hypertension, current smoking, diabetes, history of depression, sleep disturbances), are routinely captured by EHRs^12^. Recently, several studies have explored EHR data for AD/ADRD risk prediction using statistical models (e.g., the COX model^25^) and machine learning algorithms. Longitudinal patient information including diagnoses, medications, lab tests and other medical procedures were investigated as predictors. For example, Nori *et al*. developed a prediction model using an EHR-claims combined dataset and gradient boosting tree (GBT) method^26^. Park *et al*. developed AD risk prediction models with an EHR-based dataset from South Korea and explored three machine learning algorithms including logistic regression (LR), support vector machines (SVM), and random forests (RF)^27^. Both Nori *et al*. and Park *et al*. carried out the experiments in a data-driven pathway where discrete diagnoses codes, medications, and procedures were directly used to develop the models without considering clinical knowledge of AD/ADRD. They also followed a similar study design to assess the performance of predictive models using different time windows from 1-year to 4-years before the onset of AD/ADRD.

This study seeks to utilize EHRs data from the University of Florida Health (UF Health) to identify patients at risk of developing AD/ADRD before the onset of clinical symptoms and diagnosis. We explored both knowledge-driven and data-driven approaches for early prediction of AD/ADRD among elderly patients from the UF Health Integrated Data Repository (IDR). For the knowledge-driven approach, we used well-established AD/ADRD risk factors identified by our domain experts. For the data-driven approach, we directly used EHR data as features in the machine learning models. We explored early prediction using four prediction window sizes of 0-years, 1-year, 3-years and 5-years and systematically examined four widely used machine learning algorithms including LR, SVM, RF, and GBT. Our ultimate goal is to develop a robust ADRD predictive model that can be integrated into decision support systems to alert doctors in advance when their patients are at a high risk of AD/ADRD.

## Methods

### Data

In this study, we used EHR data from UF Health IDR. Supported by the UF Clinical and Translational Institute (CTSI) and UF Health, the UF Health IDR is a secure, clinical data warehouse (CDW) that aggregates data from the university’s various clinical and administrative information systems, including the Epic EHR system. From the UF Health IDR, we constructed a patient cohort using the following three inclusion criteria: 1) having at least one primary care provider visits before Jan 1st, 2014 and one after Jan 1st, 2014; 2) age ≥ 50 at the first primary care provider visit; 3) the timespan between the first and the last primary care provider visits ≥ 5 years. Since the initial documentation of AD/ADRD diagnoses are made typically by primary care physicians, we required primary care provider visits in the inclusion criteria to ensure that the patients were free of AD/ADRD diagnoses initially, to detect AD/ADRD diagnoses subsequently, and to ensure that the cohort of selected patients received outpatient services from UF Health prior to and during the study period (with respect to the documentation of risk factors for AD/ADRD). After we identified the cohort, we further collected the selected patients’ structured EHRs data including their demographics, diagnoses, medications, medical procedures, medical findings, and lab tests. This study was approved by the UF Institutional Review Board.

### Definition of cases and controls

We followed a standard case-control study design and matched controls to cases using the incidence density matching approach^28^. The cases and controls of this study were defined using the ICD-9 and ICD-10 diagnoses codes. To facilitate description, we denoted the first and last primary care encounter dates of patients as first ambulatory encounter date (FAED) and last ambulatory encounter date (LAED) in the following sections, respectively.

#### Definition for cases

Cases were defined as patients have at least one qualified ICD-9 or ICD-10 code of cognitive impairment, dementia or Alzheimer’s disease listed in Table 1. If a patient has only MCI diagnoses (ICD-9: 331.83; ICD-10: G31.84) in medical history without any other AD/ADRD diagnoses, we will exclude the patient as an invalid case.

**Table 1.** The ICD-10 and ICD-9 diagnoses codes for AD/ADRD.

| ICD-10 | Definition | ICD-9 | Definition |
| --- | --- | --- | --- |
| G30.0 | Alzheimer's disease with early onset | 290.12 | Presenile dementia with delusional features |
| G30.1 | Alzheimer's disease with late onset | 331.0 | Alzheimer's disease |
| G30.8 | Other Alzheimer's disease | 290.40 | Vascular dementia, uncomplicated |
| G30.9 | Alzheimer's disease, unspecified | 290.41 | Vascular dementia, with delirium |
| F02.80 | Dementia in other diseases classified elsewhere without behavioral disturbance | 290.42 | Vascular dementia, with delusions |
| F02.81 | Dementia in other diseases classified elsewhere with behavioral disturbance | 290.43 | Vascular dementia, with depressed mood |
| F02.90 | Unspecified dementia without behavioral disturbance | 294.0 | Amnesic disorder in conditions classified elsewhere |
| F03.91 | Unspecified dementia with behavioral disturbance | 294.8 | Other persistent mental disorders due to conditions classified elsewhere |
| F04 | Amnesic disorder due to known physiological condition | 294.9 | Unspecified persistent mental disorders due to conditions classified elsewhere |
| F06.0 | Psychotic disorder with hallucinations due to known physiological condition | 294.10 | Dementia in conditions classified elsewhere without behavioral disturbance |
| F06.8 | Other specified mental disorders due to known physiological condition | 294.11 | Dementia in conditions classified elsewhere with behavioral disturbance |
| G31.01 | Pick's disease | 294.20 | Dementia, unspecified, without behavioral disturbance |
| G31.09 | Other frontotemporal dementia | 294.21 | Dementia, unspecified, with behavioral disturbance |
| G31.1 | Senile degeneration of brain, not elsewhere classified | 331.2 | Senile degeneration of brain |
| G31.83 | Dementia with Lewy bodies | 331.6 | Corticobasal degeneration |
| G31.85 | Corticobasal degeneration | 331.7 | Cerebral degeneration in diseases classified elsewhere |
| G31.89 | Other specified degenerative diseases of nervous system | 331.11 | Pick's disease |
| G31.9 | Degenerative disease of nervous system, unspecified | 331.19 | Other frontotemporal dementia |
| G45.4 | Transient global amnesia | 331.81 | Reye's syndrome |
| G93.7 | Reye's syndrome | 331.82 | Dementia with lewy bodies |
| G94 | Other disorders of brain in diseases classified elsewhere | 331.89 | Other cerebral degeneration |
| I67.5 | Moyamoya disease | 437.7 | Transient global amnesia |
| G91.0 | Communicating hydrocephalus | 331.3 | Communicating hydrocephalus |
| G91.1 | Obstructive hydrocephalus | 331.4 | Obstructive hydrocephalus |
| G91.2 | (Idiopathic) normal pressure hydrocephalus | 331.5 | Idiopathic normal pressure hydrocephalus (INPH) |
| F01.50 | Vascular dementia without behavioral disturbance | 437.0 | Cerebral atherosclerosis |
| F01.51 | Vascular dementia with behavioral disturbance | 437.1 | Other generalized ischemic cerebrovascular disease |
| I67.1 | Cerebral aneurysm, nonruptured | 437.2 | Hypertensive encephalopathy |
| I67.2 | Cerebral atherosclerosis | 437.3 | Cerebral aneurysm, nonruptured |
| I67.4 | Hypertensive encephalopathy | 437.4 | Cerebral arteritis |
| I67.6 | Nonpyogenic thrombosis of intracranial venous system | 437.5 | Moyamoya disease |
| I67.7 | Cerebral arteritis, not elsewhere classified | 437.6 | Nonpyogenic thrombosis of intracranial venous sinus |
| I67.81 | Acute cerebrovascular insufficiency | 437.8 | Other ill-defined cerebrovascular disease |
| I67.82 | Cerebral ischemia | 437.9 | Unspecified cerebrovascular disease |
| I67.89 | Other cerebrovascular disease |  |  |
| I67.9 | Cerebrovascular disease, unspecified |  |  |

#### Definition for controls

Controls were defined as patients who have no AD/ADRD diagnoses codes listed in Table 1.

#### Case-controls matching

For each case, we matched up to nine controls according to the criteria as follows: 1) having the same gender and race as the case; 2) age within a one-year interval of the case; 3) FAED and LAED within a 30-days interval of the case; 4) having an encounter within 30-days of the case’s onset date. The encounter date closest to the onset date is denoted as the reference encounter date. After matching, we removed the cases that did not have any matched controls from the dataset. Since the prediction window size in this study is up to 5-years, we further required that the cases and controls have at least six years of EHR records. Thus, we have at least one year of data (where the prediction window = 5 years) for early prediction of AD/ADRD. Figure 1 shows an overview of the incidence-density matching procedure. Using this procedure, we identified a group of 9,414 patients, including 1,365 cases and 8,049 controls with an overall case-control matching rate of 1:5.9.

**Figure 1.**
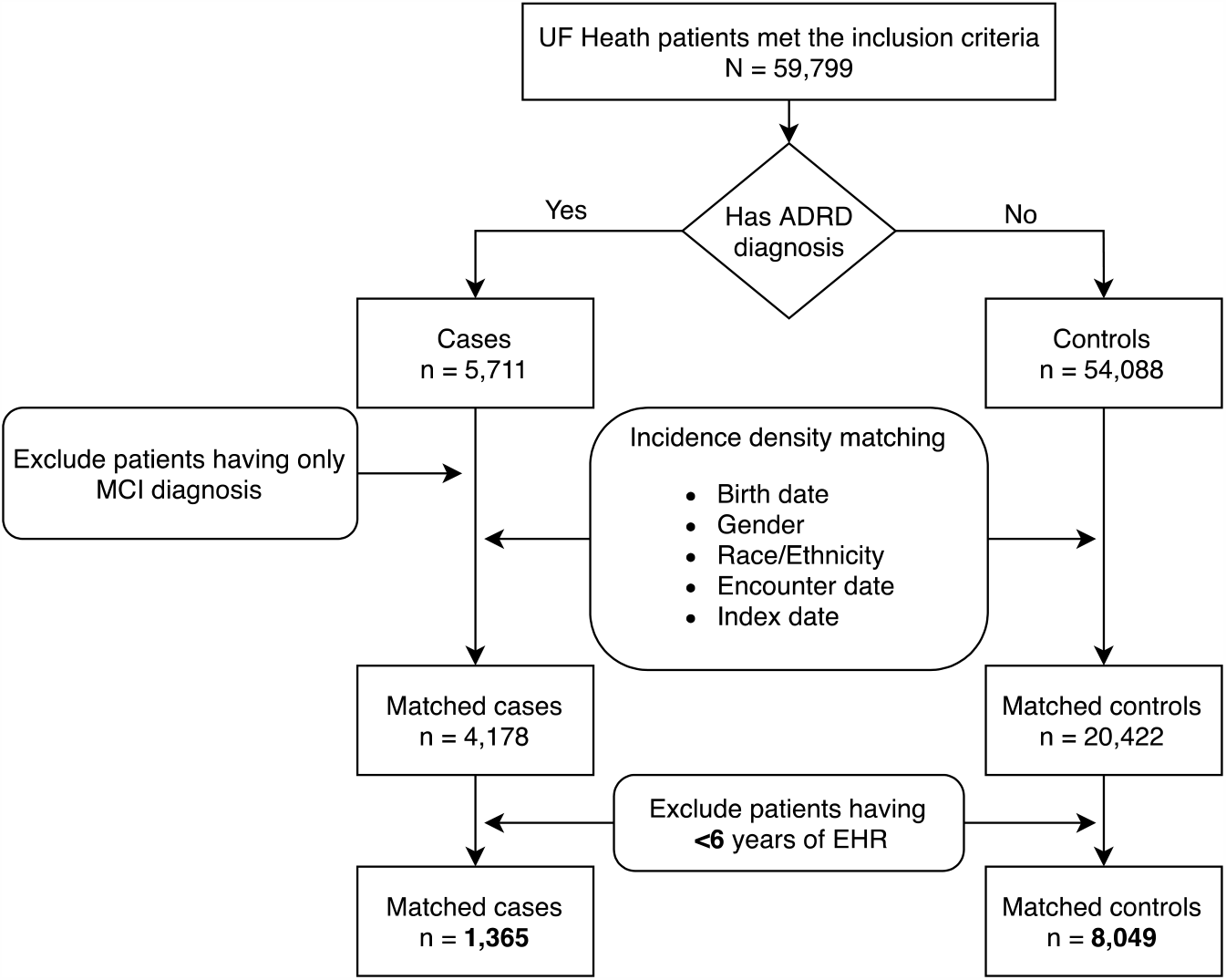
An overview of the cohort selection and case-control matching procedure.

#### Prediction window selection

This study aims to predict the risk of first onset of AD/ADRD using patients’ EHR data before the onset date of AD/ADRD. Figure 2 shows an illustration of a patient’s timeline. We utilized the patients’ data within the observation period to predict the risk of AD/ADRD after the prediction window. Here, the prediction window is defined as a censoring period from the end of observation date to the onset date of AD/ADRD. The patients’ information within the prediction window was not used (i.e., censored) for the prediction. By setting the prediction window to N-years, we were able to assess the performance of the risk prediction N-years before the onset of AD/ADRD. In this study, we systematically examined different prediction window sizes of 0, 1, 3, and 5-years. For each prediction window size, we trained an individual predictive model to optimize the features and parameters. Therefore, for each machine learning algorithm, we trained a total number of four models, including the 0-years model, 1-year model, 3-years model, and 5-years model. The 0-years model used all EHR data before the onset date of AD/ADRD (i.e., prediction window N=0).

**Figure 2.**
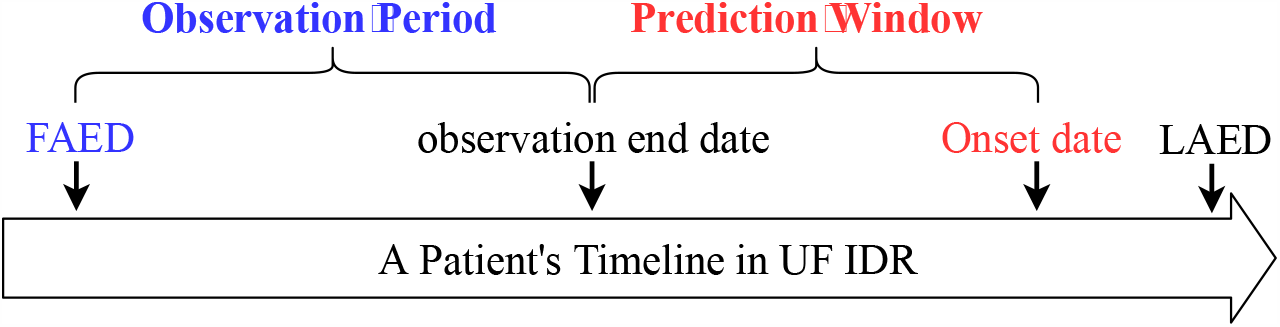
Illustration of a patient’s timeline for early prediction.

### Machine learning features

In this study, we compared two strategies to identify useful features for prediction, including a knowledge-driven approach and a data-driven approach.

#### Knowledge-driven features

For the knowledge-driven approach, we used risk factors identified by domain experts including: 1) Diagnoses of obesity, diabetes, hyperlipidemia, hypertension, heart disease, stroke, depression, anxiety, traumatic brain injury, sleep disorders, periodontitis, smoking, and alcohol use. 2) History of medication exposures to NSAIDS, statins, anticholinergics, hormone replacements, anti-hypertensive, benzodiazepines, proton pump inhibitors. 3) Latest observation of BMI, diastolic/systolic blood pressure, and lab results including total cholesterol, high density lipoprotein, fasting blood glucose, and HbA1C. More specifically, we extracted ICD-9/10 codes for the diagnoses identified by experts, RxNorm concept unique identifiers (RXCUIs) for the medications, and the Logical Observation Identifiers Names and Codes (LOINC) and values. BMI and blood pressure (systolic and diastolic) profiles were extracted from patient vitals. For the continuous data, outliers were removed based on the quantile [0.001, 0.999]; then the mean, median, minimum, maximum, standard deviation, first, and latest values within the selection were extracted for each patient.

#### Data-driven features

For the data-driven approach, we didn’t make assumptions for any risk-factors. Instead, we collected all variables captured by the EHRs (including and beyond those listed for the knowledge-driven approach) and directly used them with a minimal pre-processing in the models to let the machine learning algorithms decide the useful features for prediction. More specifically, for the demographics, we used the categorized age, gender, race, ethnicity, marital status, and smoking status. We included all discrete diagnoses codes, all medication RxNorm codes, and procedure codes as the categorical features. To reduce the sparsity of features, we adopted a similar code grouping/merging strategy described in our previous study^29^. For diagnoses, we mapped a total of 30,064 unique ICD-9/10 codes to a number of 1,912 associated PheWas (Phenome-wide association studies) groups provided by the PheWAS catalog^30^. For medications, we aggregated all the RXCUIs (22,492 unique in total) to their ingredient-level resulting in a total of 3,159 unique ingredient-level RXCUIs. We aggregated all 11,880 unique procedure codes (including CPT and ICD-9/10) into 228 unique Clinical Classification Software (CCS) groups. We encoded all the categorical features using the one-hot encoding scheme for the machine learning experiments. Real-valued variables (i.e., BMI, systolic, and diastolic) and lab tests (encoded as LOINC), were treated as continuous predictors in the model. Since a patient can have multiple findings and/or lab test measurements from different encounters, we only used the values from the most recent encounter before the observation end date. We only kept the lab tests received by more than 50% of the patients for the predictive models. For missing data, we imputed the values using a randomly selected similar sample. Finally, we further scaled the continuous variables according to the Interquartile range (IQR)^31,32^.

#### Machine learning algorithms

We systematically explored four widely used machine learning algorithms including LR, SVMs, RFs, and GBT. We applied LR as the baseline method and compared it with SVMs, RFs, and GBT. For LR, RFs, and SVMs, we adopted implementations in the Sciki-learn library^33^. For GBT, we used the implementation from the LightGBM package^34^. We optimized the hyperparameters for LR, SVMs, and RFs according to the procedures described previously^29^. For the GBT, we examined a set of hyperparameters including the number of boost trees, number of tree leaves, tree maximum depth, learning rate, and L1 and L2 regularization terms.

#### Experiments and evaluation

We split the preprocessed data into a training set of 7,487 patients (1,092 cases; 6,395 controls) and a test set of 1,927 patients (273 cases; 1,654 controls) using the stratified sampling method with the train-test ratio set as 4/1. Following best practices, we optimized all the machine learning models on the training set via five-fold cross validation and hyperparameter randomized searching. We used the area under the receiver operating characteristic curve (AUC or AUC-ROC) as the evaluation metric and reported the model performances on the test set. We also reported the sensitivity and specificity determined using the Youden’s index to facilitate model comparison^35^.

## Results

At UF Health, we identified a retrospective patient cohort of 59,799 patients from January 1, 2011 to December 31, 2018. Using incidence density sampling, the overall case-control matching ratio was 1:5.9. Table 2 compares the descriptive statistics between the case group and the control group. The distributions of the demographics (age, gender, race/ethnicity) were similar between the cases and controls. The age at index date of the case group was ∼2 years older than the control group (71.99 vs. 69.95, *p*-value<0.0001). In both case and control groups, the ratio between male and female was ∼2:3 (*p*-value =0.0539). Compared to the control group, the case group had lower percentage of Non-Hispanic White but higher percentage of other race categories (*p*-value =0.0078). The large sample size (9,414 patients in total) in this study reduced the random error and the *p*-values of the statistical tests leaned towards a small value^36^. While the cases were older and had a higher proportion of ethnic minorities (i.e., fewer non-Hispanic whites), the Cohen’s d effect sizes were small^37^ (e.g., d_age_ = 0.245, d_race_ = 0.032). We computed the medians and interquartile ranges to compare the distribution of variables between the cases and controls and found that patients in the case group had significantly more diagnoses codes, medications, procedures, medical findings and lab tests compared to patients in control group (with all *p*-values < 0.0001).

**Table 2.** Comparison of the descriptive statistics between case and control groups.

| Variable | Case<br>(N=1,365) |  | Control<br>(N=8,049) |  | P-Value |
| --- | --- | --- | --- | --- | --- |
| Demographics | N (or Mean) | % (or SD) | N (or Mean) | % (or SD) |  |
| Age at index date (year) | 71.99 | 9.03 | 69.95 | 8.14 | <0.0001<sup>a</sup> |
| Gender |  |  |  |  | 0.0539<sup>b</sup> |
| Female | 818 | 59.93 | 5,039 | 62.60 |  |
| Male | 547 | 40.07 | 3,010 | 37.40 |  |
| Race/Ethnicity |  |  |  |  | 0.0078<sup>b*</sup> |
| Non-Hispanic White | 943 | 69.08 | 5,874 | 72.70 |  |
| Non-Hispanic Black | 366 | 26.81 | 1,990 | 24.63 |  |
| Hispanic | 35 | 2.56 | 134 | 1.66 |  |
| Other | 20 | 1.47 | 30 | 0.37 |  |
| Unknown | 1 | 0.07 | 52 | 0.64 |  |
| EHR data | Median | IQR (Q1 – Q3) | Median | IQR (Q1 – Q3) |  |
| Diagnosis | 113 | 66 – 207 | 92 | 53 – 157 | <0.0001<sup>c</sup> |
| Procedures | 1,427 | 764 – 2,686 | 839 | 430 – 1,529 | <0.0001<sup>c</sup> |
| Medications | 342 | 172 – 673 | 194 | 95 – 372 | <0.0001<sup>c</sup> |
| Medical Findings | 1,339 | 627 – 3,039 | 643 | 301 – 1,337 | <0.0001<sup>c</sup> |
| Lab tests | 1,126 | 156 – 3,342 | 396 | 90 – 1,117 | <0.0001<sup>c</sup> |

a: two sample t-Test; b: Chi-square test; c: Wilcoxon rank sum test; \*: Other and Unknown were not considered in the test.

Table 3 shows the experimental results for both the knowledge-driven and data-driven approaches with different prediction windows of 0, 1, 3, and 5-years. For the 0-year model (the optimum performance we can achieve using all EHR data before the onset date), the GBT model achieved the best AUC score of 0.7976 using the data-driven approach. For 1, 3, and 5-years models, the GBT model with the data-drive approach achieved the best AUC scores of 0.7192, 0.6985, and 0.6798, respectively. Generally, the GBT model achieved the best performance for the data-driven approach, whereas, the RF model achieved the best performance for the knowledge-driven approach. For the prediction windows of 0 and 5-years, the GBT models with data-drive features are significantly better than the RF models with knowledge-driven features. However, there were no significant performance differences between the models with the knowledge-driven features and the data-driven features for the prediction window of 1, and 3-year.

**Table 3.** Comparison of the performances for 0, 1, 3, 5-years models with knowledge-driven and data-driven features.

| Prediction window | Algorithm | Knowledge-driven approach |  |  | Data-driven approach |  |  |
| --- | --- | --- | --- | --- | --- | --- | --- |
|  |  | AUC | Sensitivity | Specificity | AUC | Sensitivity | Specificity |
| 0-years | LR | 0.6866 | 0.5824 | 0.7116 | 0.7465 | 0.5421 | **0.8386** |
|  | SVM | 0.7150 | 0.6520 | 0.6826 | 0.7415 | 0.6154 | 0.7684 |
|  | RF | **0.7554** | 0.6081 | **0.7920** | 0.7855 | 0.7436 | 0.7080 |
|  | GBT | 0.7493 | **0.7399** | 0.6566 | **0.7976** | **0.8242** | 0.6566 |
| 1-year | LR | 0.6442 | 0.6117 | **0.6173** | 0.6731 | 0.6374 | 0.6330 |
|  | SVM | 0.6799 | 0.6850 | 0.5840 | 0.6821 | 0.6520 | 0.6548 |
|  | RF | **0.7104** | **0.7143** | 0.6149 | 0.7169 | 0.6886 | 0.6294 |
|  | GBT | 0.6962 | 0.7033 | 0.6028 | **0.7192** | **0.7070** | **0.6584** |
| 3-years | LR | 0.6220 | 0.5788 | 0.6505 | 0.6482 | 0.5348 | 0.7044 |
|  | SVM | 0.6486 | 0.5751 | 0.6941 | 0.6559 | **0.6557** | 0.6004 |
|  | RF | **0.6853** | **0.6923** | 0.5961 | 0.6791 | 0.5824 | 0.6820 |
|  | GBT | 0.6642 | 0.5751 | **0.7062** | **0.6985** | 0.6190 | **0.6838** |
| 5-years | LR | 0.6149 | 0.5201 | 0.6892 | 0.6259 | 0.4322 | 0.7842 |
|  | SVM | 0.6329 | 0.5385 | 0.7255 | 0.6075 | **0.7033** | 0.5067 |
|  | RF | **0.6454** | **0.6410** | 0.5852 | 0.6766 | 0.6044 | 0.6632 |
|  | GBT | 0.6389 | 0.3993 | **0.8186** | **0.6798** | 0.5568 | **0.7164** |
| Best AUCs of the knowledge driven and data driven methods for each prediction window are highlighted in bold |  |  |  |  |  |  |  |

## Discussion and Conclusion

In this study, we compared four different machine learning algorithms for early prediction of patients at risk of AD/ADRD using a cohort of 59,799 patients identified from UF Health IDR. We systematically examined the predictive models with different prediction window sizes and compared data-driven features with knowledge-driven features identified by domain experts. The experimental results show that the GBT model with data-driven features achieved the best AUC scores of 0.7976, 0.7192, 0.6985, and 0.6798 for 0, 1, 3, 5-years prediction of AD/ADRD, respectively. This study demonstrated the feasibility of using EHR data for the early prediction of patients at risk of AD/ADRD.

We compared two strategies to select useful features for the machine learning models as data-driven and knowledge-driven approaches. For the data-driven approach, the machine learning models selected useful features among all patient’s information available in the EHRs. On the other hand, for the knowledge-driven approach, our domain experts identified the top features that highly associated with the risk of AD/ADRD. For the data-driven approach, the GBT model achieved the best AUC score outperforming other machine learning methods for 0, 1, 3, 5-years prediction windows. For the knowledge-driven approach, the RF model achieved the best performances among all models explored. For both approaches, the performances decreased when increasing the size of the prediction window. For example, the performance of the GBT model with the data-driven approach remarkably dropped from 0.7976 to 0.7192 when the prediction window increased from 0-years to 1-year. Similarly, when further increasing the prediction window to 3-years and 5-years, the AUCs consistently dropped to 0.6985 and 0.6798. This result is not surprising as the observation period used for prediction becomes shorter when increasing the prediction window size (see Figure 2). Consequently, we have fewer features extracted from the EHR data for building the prediction models. Taking the data-driven approach as an example, the total number of features were 7,766, 7,282, 6,027, and 4,669 for the 0, 1, 3, 5-years models, respectively. Therefore, the prediction performance dropped as we were losing patients’ information by increasing the prediction window.

The experimental results showed that the data-driven approach was able to utilize more predictive features from EHRs for early prediction of patients at risk of ADRD compared with the knowledge-driven approach. Therefore, the data-driven based models had better predictive power, especially for the 0-years and 5-years models. For example, in the 5-year prediction, the GB model with the data-driven approach achieved the best AUC of 0.6798, outperforming the knowledge-driven model of 0.6454. Similar results were observed for 0-, 1-, and 3-years predictions as well. We further extracted the top 100 features from the GBT models with the data-driven approach based on feature importance. Among these features, most of the knowledge-driven features identified by domain experts were captured, such as age, blood pressure (systolic and diastolic), diabetes, stroke, heart disease, depression, and cholesterol. However, there were a few knowledge-driven risk factors such as anxiety, sleep disorder, and periodontitis that were not captured by the data-driven approach. In addition, the data-driven approach identified new essential risk factors, including mean arterial pressure, seizures, hemoglobin, and marital status. We further added the newly discovered risk factors identified by the data-driven model to the knowledge-driven features and retrained an RF model – the best machine learning model for the knowledge-driven approach. The preliminary results showed that the new features further improved the performance of the knowledge-driven approach. For example, for the 5-years prediction, the performance was improved from 0.6454 to 0.6589. Although many features identified in the data-driven approach were clinically meaningful risk factors, further investigations are needed to examine the new features that were not identified in previous AD/ADRD prediction research, such as diagnoses of dry eyes, hearing loss, viral hepatitis C, and the procedure of routine chest X-ray.

Our experimental results are similar to the previous studies of exploring EHR data for early prediction of AD/ADRD. In studies from Nori *et al*.^26^ and Park *et al*.^27^, the AUC scores decreased with the increase of the prediction window sizes. For example, Park and co-workers showed that expanding the prediction window from 0-years to 4-years, the AUC score achieved by the best machine learning model (RF-based) dropped remarkably from 0.805 to 0.602. In both our models and the models developed by *Nori et al*., Parkinson’s disease, depression, and CT scan head were identified as top important features for prediction. Both our models and the models by Park *et al*. identified hemoglobin as an important predictor. In addition to the well-known risk factors, our study further exhibited that chronic pain is an important feature not reported in the previous two studies. Our discovery is consistent with the a recent clinical study reporting a strong association between chronic pain and dementia among elderly population^38^.

This study has limitations. The UF Health IDR started tracking patients’ records in 2011. Therefore, our current work may suffer from incomplete data, varying length of observation, and coding bias as we cannot access patients’ information before 2011. We mainly focused on traditional machine learning models that could not utilize the time sequence information associated with the clinical variables (e.g., diagnoses, medications). We combined AD and ADRD in one category for prediction and did not consider their subtypes (e.g., vascular dementia or Lewy body dementia vs. AD). Our future work seeks to examine social and behavioral determinants of health information documented in the narrative clinical notes (e.g., education) and explore time-to-event prediction models that could utilize the time sequence information such as recurrent neural networks-based models (e.g., RETAIN^39^).

This study investigated early prediction of patients at risk of AD/ADRD using EHR data from the UF Health. We compared a knowledge-driven approach with a data-driven approach, and systematically examined four widely-used machine learning models. Our study demonstrated the feasibility of using EHR for early prediction of AD/ADRD and discovered potential challenges. Compared with early prediction of other diseases (e.g., our previous work on heart failures^29^), further investigations are needed to improve the prediction of AD/ADRD.

## Data Availability

Data sharing is not applicable to this article.

## Acknowledgement

This study was partially supported by an Ed and Ethel Moore Alzheimer’s Disease Research Program from the Florida Department of Health (FL DOH #9AZ14) and a Patient-Centered Outcomes Research Institute® (PCORI®) Award (ME-2018C3-14754). The content is solely the responsibility of the authors and does not necessarily represent the official views of the funding institutions.

